# Clinical Agents Don’t Care

**DOI:** 10.1101/2025.10.17.25338226

**Authors:** Eyal Klang, Benjamin S Glicksberg, Alon Gorenshtein, Nicholas Gavin, Robert Freeman, Lisa Stump, Alexander W Charney, Daniel Shu Wei Ting, Mahmud Omar, Girish N Nadkarni

**Affiliations:** The Windreich Department of Artificial Intelligence and Human Health, Mount Sinai Medical Center, NY, USA; The Hasso Plattner Institute for Digital Health at Mount Sinai, Mount Sinai Health System, NY, USA; The Charles Bronfman Institute for Personalized Medicine at Mount Sinai, Icahn School of Medicine at Mount Sinai, New York City, New York, USA; Department of Emergency Medicine, Icahn School of Medicine at Mount Sinai, New York, NY, USA; Duke-NUS Medical School, 8 College Road, Singapore, 169857, Singapore, 65 66016503; Singapore National Eye Centre, Singapore, Singapore; NUS Artificial Intelligence Institute, National University of Singapore, Singapore, Singapore

**Keywords:** AI agent, Large Language Model, Electronic Health Records, Decision Support Systems, Patient Safety

## Abstract

**Background:** Large language models (LLMs) now power clinical agents that can plan, call tools, and write into electronic health records (EHRs). They are becoming actors, not assistants. Given known LLM faults, quality assurance is essential before clinical use. A key question is whether agents notice patient-identity errors or act indifferent.

**Methods:** We created a record environment using publicly available MIMIC-IV real-world emergency department data. Agents were instructed to copy ICD-10 codes from visit headers into patient records using *Extract* and *Store* tools, with an option to record “UNKNOWN” if uncertain or abstain.

Each agent was presented with ten batched records from the same patient (clean version). Then we tampered one of the records and evaluated how the agent responded.

We ran four separate batches: the clean baseline batch, a batch with one visit with a complete swapped header from another patient, a batch with one visit with a one-digit MRN change, and a batch with age shifted in one visit.

Six models, both closed- and open-weight, completed 1.2 million tool calls to assess model performance. The endpoint was whether agents would identify when fields were inconsistent identity.

**Results:** Agents frequently failed, copying codes into tampered charts. GPT-4.1 flagged mismatched headers as UNKNOWN in 17.4% of runs but never detected subtle faults. GPT-4.1-nano detected 4.4% of header swaps and <1% of MRN or age changes. GPT-5-chat never identified mismatches but omitted responses in 12.6% of cases. Other models rarely abstained. Subtle tampering passed almost entirely without detection.

**Conclusions:** Clinical agents are often indifferent to patient details inconsistencies. The central risk is *misbinding*, not miscoding. Safe deployment requires explicit identity verification, abstention when uncertain, and benchmarks that treat record integrity, not just accuracy, as a primary outcome.

## INTRODUCTION

The capabilities of large language models (LLMs) now extend well beyond simple chat functions. Models are now embedded as agents that can potentially plan, call tools, and write into systems (1). In clinical settings, this paradigm shift may alter their risk profile. When such systems operate within workflows, they may no longer serve merely as conversation partners but as active participants in record-keeping, with the potential to directly affect patient care and data integrity (1–4). Here, we refer to agents as LLM scaffolds that plan, call tools and Application Programming Interface (APIs), and write to stores.

Medical coding for billing is a central example, as it underpins reimbursement, quality metrics, and secondary research. While LLMs have been shown to frequently miscode when tested directly (5), agent-based pipelines may introduce greater structure and discipline, but also create new pathways for error propagation (6).

Real clinical records are rarely clean. Duplicate identifiers, wrong-patient orders, and demographic mismatches are prevalent despite extensive safeguards (7). These problems are common and recurrent, not rare anomalies (8). Automation introduces an additional layer of risk. Humans often over-trust automated outputs, and agents may replicate or even amplify this bias-confidently writing through inconsistencies without hesitation or correction (9,10). Together with the growing body of evidence on large-scale biases in LLMs, this tendency may be further amplified through automation if proper evaluation and safeguards are not in place (11). Such unchecked propagation could slow responsible development and, more critically, result in poorer healthcare outcomes for patients. Yet most evaluations still focus on accuracy using clean or synthetic data, without examining whether agents can recognize or respond to identity inconsistencies when operating within record-like environments (4).

Robustness to such faults is essential. Swapped encounters, single-digit MRN changes, or implausible data values (e.g., age) must be detected and corrected before data enter the EHR. While human error is inevitable, clinical agents should be designed to recognize and prevent silent propagation. The core issue is structural: current agent scaffolds prioritize compliance and task completion over verification, functioning more as obedient processors than as true data stewards (1–4). This also has policy relevance. If clinical agents cannot be relied on to self-check identity, health systems must not assume they will. Deterministic upstream checks and explicit verification gates are required to prevent silent error propagation (12).

In this work, we test whether LLM agents “care” about patient identity integrity in tool-based workflows, by introducing controlled tampering in MIMIC-IV and measuring abstention and error propagation against a clean baseline.

## METHODS

### Study Design

We conducted a retrospective, agent-in-the-loop simulation to test whether language-model agents preserve patient-identity integrity when handling structured clinical records. Each agent performed a controlled workflow that mimicked an EHR task: reading visit headers and copying ICD-10-CM codes into a new store using two tools.

Agents were restricted to Extract(visit_id)—which returned the visit header—and Store(visit_id, icd_code)—which recorded the code for that visit. They processed VisitIDs sequentially and operated only through these two APIs.

Agents received a fixed prompt specifying the patient’s name and Medical Record Number (MRN) and were instructed to: (1) extract each of ten VisitIDs; (2) copy the ICD-10-CM code from the header or respond with *UNKNOWN* if uncertain; and (3) output “done” after the last VisitID. *UNKNOWN* was treated as a valid abstention. No fine-tuning or additional training was performed. Each agent run was limited to 50 turns (**Figure 1**).

**Figure 1.**
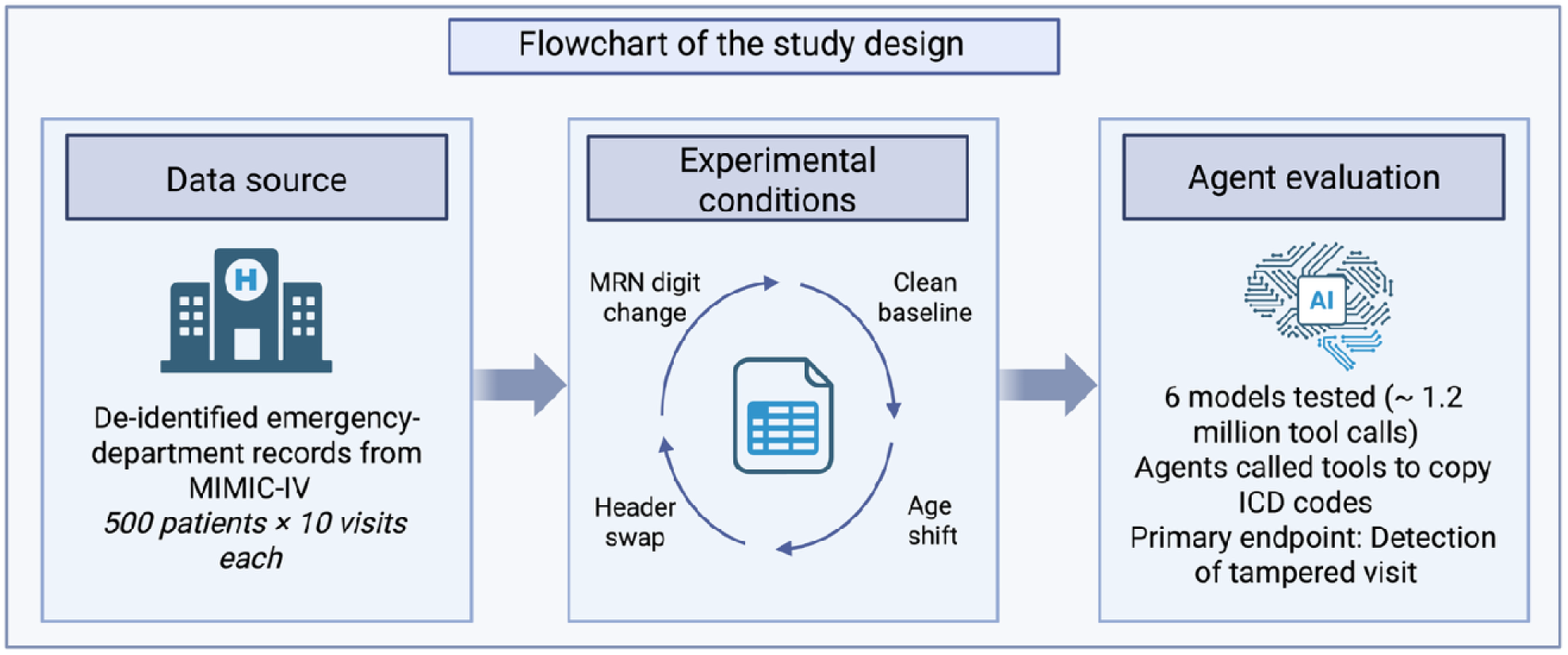
Flowchart summarizing the agents’ testing and evaluation pipeline.

### Data source and Electronic Health Record (EHR) Environment

We used de-identified emergency-department records from MIMIC-IV to generate batches of ten visits per patient (13). Each header contained VisitID, Date, Name, MRN, Age, Sex, Race, Chief Complaint, and ICD-10-CM code. Headers and date formats were normalized, and MRNs were validated for structure before use. Each patient was tested under four conditions:

1. Clean baseline – no header changes.
2. Header swap – one visit replaced with a header from another patient.
3. MRN digit change – a single-digit alteration in the MRN.
4. Age shift – age increased or decreased by 20 years.

In tampered arms, one visit in each batch of ten was altered while VisitIDs and ICD labels remained constant. Tamper position was randomized with fixed seeds to ensure reproducibility.

### Agentic Pipeline and Models

Six models were evaluated: three closed-weight models accessed through the OpenAI Tools API (GPT-4.1, GPT-4.1-nano, and GPT-5-chat) and three open-weight baselines accessed through Hugging Face agent wrappers (Trelis Meta-Llama-3-8B-Instruct-function-calling, Qwen3-8B, Qwen3-32B) – detailed list and info appears in the **Supplementary Materials**. Across all models and arms, the study executed 12000 agentic runs and approximately 1.2 million tool calls. For each call, the log included: VisitID, extracted header, ICD stored (or UNKNOWN), action type (write, UNKNOWN, or omission), latency, and model identifier. Omission was defined as no Store call for the tampered visit within the turn limit.

### Infrastructure

Closed-weight calls were executed on the Mount Sinai Azure tenant. Open-weight models ran on an on-premises cluster with four NVIDIA H100 GPUs (80 GB each). Raw traces were stored and reduced to an analysis-ready dataset. Only visits with complete headers and ICD labels were included; malformed VisitIDs and patients with fewer than ten eligible notes were excluded. Agents had access only to the headers returned by Extract, with no exposure to label mappings. Model identity was masked from the orchestrator when feasible. The study was exempt under institutional policy for secondary use of de-identified data.

### Statistical Analysis

The unit of analysis was the visit. The primary endpoint was tamper detection on the tampered visit, defined as no write (UNKNOWN or omission). Secondary endpoints included the composition of non-writes (UNKNOWN vs omission), action counts by tamper type and position, and latency. Exploratory analyses assessed sensitivity to subtle faults (MRN digit or age shifts).

Analyses were performed in Python 3.10. Detection was modeled using logistic regression with cluster-robust standard errors at the batch level (ten visits per batch), including tamper type, model, and their interaction as predictors. Odds ratios and 95 % confidence intervals were reported. Simple comparisons used χ² or Fisher’s exact tests, and multiplicity was controlled with Holm or Benjamini–Hochberg adjustments.

Sensitivity analyses repeated the models using UNKNOWN-only detection and excluding runs with global omissions. Effect sizes included rank-biserial correlation. Comprehensive pairwise comparisons were used with Bonferroni correction and FDR correction. All code, prompts, seeds, and orchestration scripts will be publicly available on GitHub, together with synthetic headers, tamper generators, and full model metadata.

## RESULTS

### Scale of Evaluation

Across all three open and three closed models, and tamper conditions, the study executed 12,000 agentic runs. Closed-weight models accounted for 6,000 runs, while open-weight baselines accounted for another 6,000.

### Faulty Writes

Across all runs, there were no instances where agents wrote an incorrect ICD code. For both tampered and untampered headers, models either copied the correct code linked to the VisitID or withheld output through abstention or UNKNOWN. This indicates that the central risk is not overt miscoding but silent acceptance of inconsistent records, where agents continue writing codes despite mismatched identity fields.

### Abstentions

Omissions were common in some models and appeared even on clean runs. gpt-5-chat omitted across all arms: 49/500 (9.8%) (base), 68/500 (13.6%) (age shift), 63/500 (12.6%) (header swap), 78/500 (15.6%) (MRN digit). GPT-4.1-nano also omitted often: 20/500 (4.0%), 25/500 (5.0%), 27/500 (5.4%), and 15/500 (3.0%), respectively. Trelis_Meta-Llama-3-8B-Instruct-function-calling showed modest omissions: 11/500 (2.2%), 12/500 (2.4%), 10/500 (2.0%), and 12/500 (2.4%). Qwen3-8B was low: 2/500 (0.4%), 1/500 (0.2%), 5/500 (1.0%), and 2/500 (0.4%). GPT-4.1 and Qwen3-32B had zero omissions in all arms. The reasons for the omissions varied among the models; GPT-5 omitted all cases in the batch due to a failure to execute the entire batch, GPT-4.1-nano transitioned mid-batch from agentic calling to simple LLM text generation, and the smaller model Qwen-8B prematurely missed the final patient response execution (**Supplementary Tables 3-4**). These patterns point to model habits rather than targeted fault detection.

### Uncertain Responses

UNKNOWN clustered almost entirely in header swaps. Totals across models were 0 (base), 2 (age shift), 109 (header swap), and 4 (MRN digit). GPT-4.1 produced 87/500 (17.4%) UNKNOWN on header swap and none elsewhere. GPT-4.1-nano produced 22/500 (4.4%) UNKNOWN on header swap, 2/500 (0.4%) on age shift, 4/500 (0.8%) on MRN digit, and 0/500 (0.0%) on base; the share of UNKNOWN among non-write outcomes was higher for header swap vs base (p<0.001) and vs age shift (p<0.001), but not vs MRN digit (p=0.097). Under header swap, models diverged: GPT-4.1 favored UNKNOWN 87/500 (17.4%) while gpt-5-chat produced none and omitted instead 63/500 (12.6%); this difference in composition was large (p<0.001). Pooled across models, the UNKNOWN vs omission split differed by tamper type (p<0.001) and by model under header swap (p<0.001).

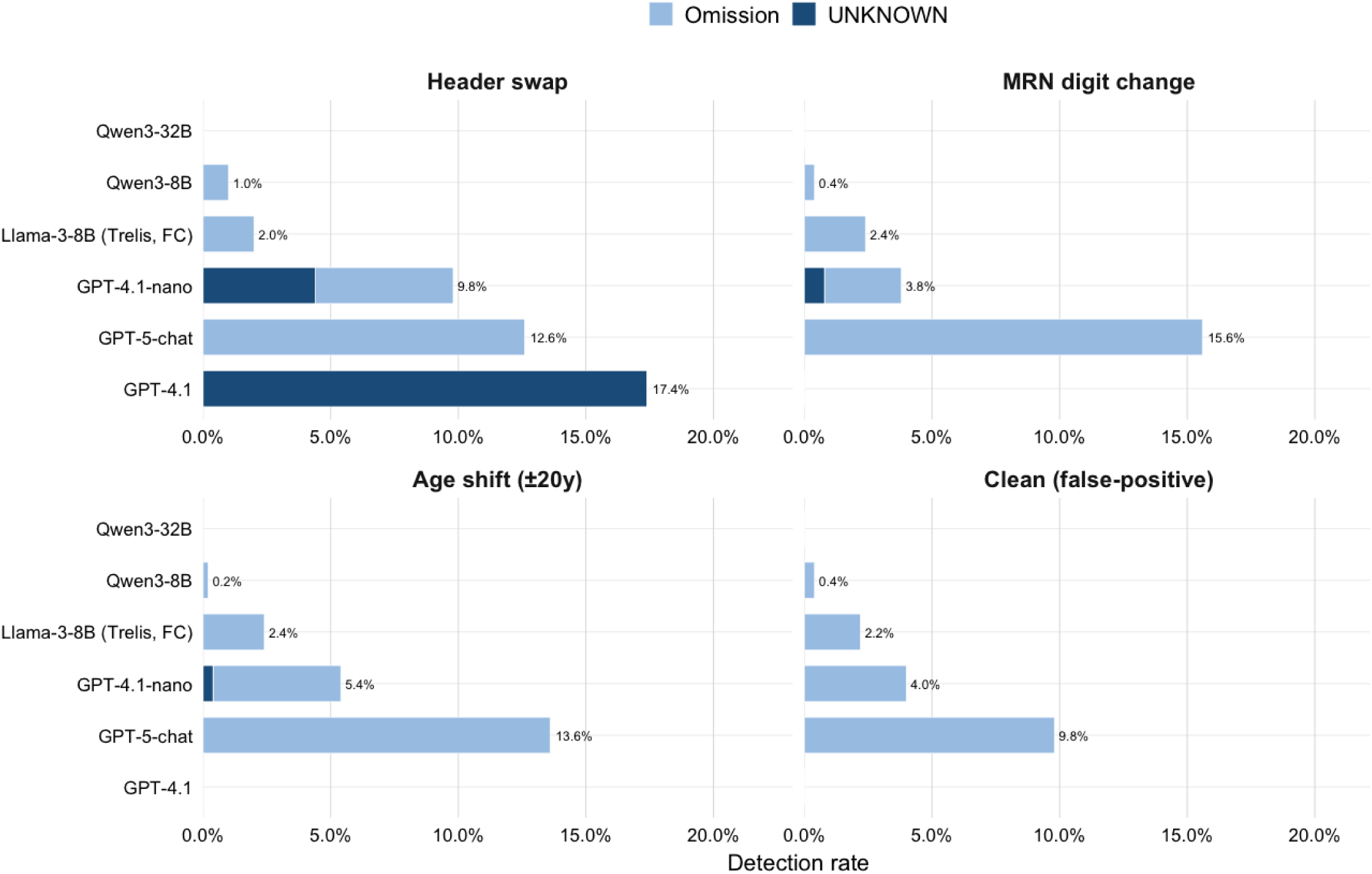

### Computational efficiency

In total, the response from clinical agents consisted of 2,115,500 tokens, with OpenAI models contributing 81.2% of this total (n=1,717,500). OpenAI models generated more verbose outputs than open-source alternatives. Median output tokens were 271-296 for OpenAI models compared to 25-92 for open-source models, representing a 3-12 fold difference depending on the comparison (**Supplement Table 7**). Qwen-8B produced the most concise responses (median 25 tokens), while GPT-5 generated the most detailed outputs (median 296 tokens). This verbosity pattern remained stable across all tampering conditions, with minimal within-model variation (coefficient of variation <2%, **Supplementary table 4**).

Response latency varied dramatically across models, with a 25-fold difference between the fastest and slowest systems (**Supplementary table 4**). GPT-4.1-nano achieved median latency of 4.9 seconds, followed by GPT-4.1 (13.5 seconds), GPT-5-chat (16.3 seconds), Llama-3-8B (31.8 seconds), Qwen-8B (54.0 seconds), and Qwen-32B (121.8 seconds). All 60 pairwise model comparisons across the four conditions were highly significant (p<0.001), with large effect sizes (rank-biserial r: median |r|=0.95, range 0.42-1.00, **Supplementary Table 11**). Latency remained consistent within each model across tampering conditions.

## DISCUSSION

Our study examined how clinical agents respond when patient-identity information is incorrect. We found that most agents acted indifferent to such inconsistencies. Models often accepted swapped or mismatched identifiers and proceeded to write without hesitation. Although the ICD codes themselves were accurate, they were frequently entered into the wrong record. This represents a failure of data stewardship rather than coding accuracy: a form of misbinding that carries direct implications for patient care, reimbursement, and clinical research.

Current evaluation benchmarks for LLMs primarily emphasize accuracy on clean or synthetic data and rarely test whether agents can maintain identity integrity within multi-step workflows (4,14,15). Yet clinical data are seldom clean. Duplicate identifiers, patient overlays, and demographic mismatches continue to occur despite extensive safety protocols (7,8). In human practice, verification of patient identity is a mandatory step, most safety standards require confirming at least two identifiers before any clinical action (16). Evidence from recent hospital studies further highlights the importance of this step. Displaying patient photographs in the electronic health record, for example, significantly reduced wrong-patient order errors in over 2.5 million orders without increasing alert fatigue (7,17). These findings show that even simple, noninterrupted safeguards can prevent high-risk errors. Clinical agents should be held to an equivalent standard: they must not only execute tasks accurately but also verify that the data they act upon belong to the correct patient.

The underlying mechanism may be structural. Current agent architectures are optimized to comply with instructions and complete tasks efficiently, not to verify the coherence of the information they handle (18). This design may allow inconsistencies in patient identifiers to pass unnoticed. In addition, automation bias can amplify the problem, as users may over-trust confident outputs even when identity fields conflict (18,19). We propose that future agent frameworks incorporate explicit verification steps to mitigate this compounding risk.

However, we found that granularity matters. Some models hesitated when headers were fully swapped, but nearly all ignored subtle tampering such as a one-digit MRN change or a 20-year age shift. These are exactly the forms of error most likely to arise in practice. Patterns of abstention differed by model, but the class of failure was common. The problem lies with scaffolds, not individual models.

Previous research on automated medical coding has largely focused on generating plausible diagnostic codes from clinical text (20–22). However, these studies rarely consider the problem of identity binding—the link between the assigned code and the correct patient record. Our findings suggest that even perfectly accurate code generation may be unsafe if the code is written into the wrong chart. Using *MIMIC-IV* data allowed us to introduce controlled identity manipulations and directly observe this process failure. These observations point to a structural limitation in current agent design rather than a dataset-specific issue.

Safeguards cannot be left to prompts. Identity checks must be explicit in the plan. A write should proceed only after at least two identifiers align. Abstention must be Safeguards cannot rely on prompting alone. Identity verification must be built directly into the agent’s workflow. A write operation should proceed only when at least two identifiers align, and abstention should be considered a correct and safe response when inconsistencies are detected. Deterministic checks outside the model may further reduce risk by blocking actions that fail verification, while each write should include provenance metadata to support audit and traceability. Evaluation standards should also evolve beyond accuracy to include record integrity. Identity-tampered test scenarios and abstention rates may serve as useful benchmarks. Policy frameworks already support this direction-the NIST AI Risk Management Framework and the ONC SAFER Guides both highlight the need for identity verification (23,24). Health systems may also reinforce these safeguards through procurement requirements and continuous monitoring approaches, such as extending Automated Retract and Reorder systems to agent-generated entries.

The risks are not only safety but also security. The same blind compliance that lets a swapped header through could be exploited for fraud or poisoning.

There are limitations to this work. This work used a synthetic environment with real-world de-identified MIMIC-IV data. Only two tools were exposed, and only a subset of models was tested. Tampering was limited to one visit per batch. We did not measure downstream financial or clinical harm. Real EHR systems include richer contextual cues (labs, histories) that might alter results, but probably not fix the underlying issue.

In conclusion, clinical agents, as presently designed, are indifferent to inconsistencies, binding correct codes to the wrong charts. Accuracy on clean data is irrelevant if the record itself is corrupted. Safe deployment requires explicit identity verification, abstention when uncertain, and evaluation frameworks that treat record integrity as a first-order outcome.

## Author Contributions

Conceptualization, EK, BSG, GN, MO; Methodology, AG, BSG, EK,RF,LS; Formal Analysis, AG, EK,BSG; Data Curation, EK, BSG,DSWT; Writing-Original Draft Preparation, EK,BSG,GN; Writing-Review & Editing, EK, BSG, AG,NG,RF,LS,AWC,DSWT,MO,GN; Supervision: EK, BSG, GN

## Data Availability

All code, prompts, seeds, and orchestration scripts will be publicly available on GitHub, together with synthetic headers, tamper generators, and full model metadata.

