## Appendix for "Clinical Agents Don’t Care"

#### Table of Contents

*Supplementary Appendix* ____ 1

**Supplementary Results** ____ 2

**Supplementary Tables** ____ 2

**Supplementary Table 1** Model specifications for all clinical agents evaluated ____ 2

**Supplementary Table 2** Counts of abstentions across all models and tamper conditions. ____ 3

**Supplementary Table 3 Failure Mechanisms** ____ 4

**Supplementary Table 4 Exact Failure Mechanisms** ____ 5

**Supplementary Table 5 Unknown count pairwise comparison of different perturbations** ____ 6

**Supplementary Table 6 Unknown count pairwise comparison of different LLMs** ____ 7

**Supplementary Table 7 Token count per LLM model** ____ 11

**Supplementary Table 8. Token count pairwise comparison of different perturbations** ____ 12

**Supplementary Table 9. Token count pairwise comparison across different LLMs** ____ 13

**Supplementary Table 10. Latency count per LLMs** ____ 17

**Supplementary Table 11. Latency count pairwise comparison across different LLMs** ____ 18

Division of Data-Driven and Digital Medicine (D3M), Icahn School of Medicine at Mount Sinai, New York, USA

Mount Sinai

### Supplementary Results

#### Supplementary Tables

**Supplementary Table 1** Model specifications for all clinical agents evaluated

| Model | Provider | Weights | Parameters | Context window | License |
|---|---|---|---|---|---|
| **GPT-4.1** | OpenAI | Closed | Undisclosed | 1M tokens | Proprietary |
| **GPT-4.1-nano** | OpenAI | Closed | Undisclosed | 1M tokens | Proprietary |
| **GPT-5-chat** | OpenAI | Closed | Undisclosed | 400K tokens | Proprietary |
| **Trelis Meta-Llama-3-8B-Instruct-function-calling** | Trelis (fine-tune) / Meta (base) | Open-weight | 8B | 8k tokens | Llama 3 Community License |
| **Qwen3-8B** | Alibaba/Qwen team | Open-weight | 8.2B | 32k tokens | Apache-2.0 |
| **Qwen3-32B** | Alibaba/Qwen team | Open-weight | 32.8B | 32k tokens | Apache-2.0 |

**Supplementary Table 2** Counts of abstentions across all models and tamper conditions.

| Model | Base U | Base O | Age shift U | Age shift O | Header swap U | Header swap O | MRN digit U | MRN digit O |
|---|---|---|---|---|---|---|---|---|
| **gpt-4.1-nano** | 0 | 20 | 2 | 25 | 22 | 27 | 4 | 15 |
| **gpt-4.1** | 0 | 0 | 0 | 0 | 87 | 0 | 0 | 0 |
| **gpt-5-chat** | 0 | 49 | 0 | 68 | 0 | 63 | 0 | 78 |
| **Trelis_Meta-Llama-3-8B-Instruct-function-calling** | 0 | 11 | 0 | 12 | 0 | 10 | 0 | 12 |
| **Qwen3-8B** | 0 | 2 | 0 | 1 | 0 | 5 | 0 | 2 |
| **Qwen3-32B** | 0 | 0 | 0 | 0 | 0 | 0 | 0 | 0 |

"U" denotes *UNKNOWN* responses, where the agent explicitly declined to write due to uncertainty. "O" denotes *omissions*, where no write occurred within the turn limit. Each column shows the number of UNKNOWN or omission outcomes under baseline, age shift, header swap, and MRN digit–change conditions.

**Supplementary Table 3** Failure Mechanisms

| Model | Omissions count | Omissions rate | Failure Stage | Workflow Stage at Failure | Data Recovery Potential | Technical Failure Mechanism |
|---|---|---|---|---|---|---|
| **GPT-4.1** | 0 | 0.0 | None | N/A | N/A | None |
| **GPT-4.1-nano** | 87 | 4.3% | Post-extraction | Post-extraction phase | None - complete loss | Function invocation failure after successful data extraction |
| **GPT-5** | 259 | 12.9% | Pre-execution | Pre-execution phase | None - no data generated | Infrastructure failure preventing workflow initialization |
| **Llama-3-8B** | 74 | 3.7% | Bimodal: Pre-execution (48.6%) and Near-completion (51.4%) | Bimodal distribution | Variable by timing | Mixed: initialization failures (48.6%) and output validation failures (51.4%) |
| **Qwen-8B** | 27 | 1.4% | Near-completion | Near-completion phase | High - 9/10 diagnoses preserved | Premature termination with off-by-one counting error |
| **Qwen-32B** | 0 | 0.0 | None | N/A | N/A | None |

**Supplementary Table 4** Exact Failure Mechanisms

| Model | Timing Category | Percent Complete | Extract Calls Made | Extract Success | Store Calls Made | Store Success | Exact Problem | Data Preserved |
|---|---|---|---|---|---|---|---|---|
| GPT-4.1 | Complete | 100% | 10 | 10/10 | 10 | 10/10 | None | 100% |
| GPT-4.1-nano | Mid (50%) | 48% | 10 | 10/10 | 0 | 0/10 | Function→Text mode switch | 0% (data lost) |
| GPT-5 | Early (0%) | 0% | 0 | 0/10 | 0 | 0/10 | Infrastructure crash | 0% (never started) |
| Llama-3-8B (Early) | Early (0-24%) | 0-24% | 0-5 | Varies | 0-5 | Varies | Infrastructure/ parsing | Varies |
| Llama-3-8B (Late) | Late (76-95%) | 76-95% | 8-10 | Varies | 8-10 | Varies | Output formatting | 80-95% |
| Qwen-8B | Late (90%) | 90% | 9 | 9/9 | 9 | 9/9 | Off-by-one counting error | 90% (9/10 codes) |
| Qwen-32B | Complete | 100% | 10 | 10/10 | 10 | 10/10 | None | 100% |

**Supplementary Table 5** Unknown count pairwise comparison of different perturbations

| Model | Comparison | Base UNKNOWN Rate | Tamper UNKNOWN Rate | Odds Ratio | p-value | Effect-Size |
|---|---|---|---|---|---|---|
| GPT-4.1 | Age shift vs Base | 0.0 | 0.0 | 1 | p=1.00 | Negligible |
| GPT-4.1 | Header swap vs Base | 0.0 | 17.4% | inf | **p<0.001** | Large |
| GPT-4.1 | MRN digit change vs Base | 0.0 | 0.0 | 1 | p=1.000 | Negligible |
| GPT-4.1-nano | Age shift vs Base | 1.2% | 1.8% | 0.663 | p=0.604 | Negligible |
| GPT-4.1-nano | Header swap vs Base | 1.2% | 5.6% | 0.205 | **p<0.001** | Small |
| GPT-4.1-nano | MRN digit change vs Base | 1.2% | 2.0% | 0.595 | p=0.45 | Negligible |
| GPT-5 | Age shift vs Base | 0.0 | 0.0 | 1 | p=1.00 | Negligible |
| GPT-5 | Header swap vs Base | 0.0 | 0.0 | 1 | p=1.00 | Negligible |
| GPT-5 | MRN digit change vs Base | 0.0 | 0.0 | 1 | p=1.00 | Negligible |
| Llama-3-8B | Age shift vs Base | 0.0 | 0.0 | 1 | p=1.00 | Negligible |
| Llama-3-8B | Header swap vs Base | 0.0 | 0.0 | 1 | p=1.00 | Negligible |
| Llama-3-8B | MRN digit change vs Base | 0.0 | 0.0 | 1 | p=1.00 | Negligible |
| Qwen-8B | Age shift vs Base | 0.2% | 0.2% | 1 | p=1.00 | Negligible |
| Qwen-8B | Header swap vs Base | 0.2% | 0.2% | 1 | p=1.00 | Negligible |
| Qwen-8B | MRN digit change vs Base | 0.2% | 0.2% | 1 | p=1.00 | Negligible |
| Qwen-32B | Age shift vs Base | 0.0 | 0.0 | 1 | p=1.00 | Negligible |
| Qwen-32B | Header swap vs Base | 0.0 | 0.0 | 1 | p=1.00 | Negligible |
| Qwen-32B | MRN digit change vs Base | 0.0 | 0.0 | 1 | p=1.00 | Negligible |

**Supplementary Table 6** Unknown count pairwise comparison of different LLMs

| Condition | Model 1 | Model 2 | Model 1 UNKNOWN Rate (%) | Model 2 UNKNOWN Rate (%) | Rank Biserial r | Odds Ratio | p-value |
|---|---|---|---|---|---|---|---|
| Base | GPT-4.1 | GPT-4.1-nano | 0.0 | 1.2 | 1.0 | 0.0 | **p=0.031** |
| Base | GPT-4.1 | GPT-5 | 0.0 | 0.0 | 0.0 | 1 | p=1.00 |
| Base | GPT-4.1 | Llama-3-8B | 0.0 | 0.0 | 0.0 | 1 | p=1.00 |
| Base | GPT-4.1 | Qwen-8B | 0.0 | 0.2 | 1.0 | 0.0 | p=1.00 |
| Base | GPT-4.1 | Qwen-32B | 0.0 | 0.0 | 0.0 | 1 | p=1.00 |
| Base | GPT-4.1-nano | GPT-5 | 1.2 | 0.0 | -1.0 | inf | **p=0.031** |
| Base | GPT-4.1-nano | Llama-3-8B | 1.2 | 0.0 | -1.0 | inf | **p=0.031** |
| Base | GPT-4.1-nano | Qwen-8B | 1.2 | 0.2 | -0.833 | 6.061 | p=0.12 |
| Base | GPT-4.1-nano | Qwen-32B | 1.2 | 0.0 | -1.0 | inf | p**=0.031** |
| Base | GPT-5 | Llama-3-8B | 0.0 | 0.0 | 0.0 | 1 | p=1.00 |
| Base | GPT-5 | Qwen-8B | 0.0 | 0.2 | 1.0 | 0.0 | p=1.00 |
| Base | GPT-5 | Qwen-32B | 0.0 | 0.0 | 0.0 | 1 | p=1.00 |
| Base | Llama-3-8B | Qwen-8B | 0.0 | 0.2 | 1.0 | 0.0 | p=1.00 |
| Base | Llama-3-8B | Qwen-32B | 0.0 | 0.0 | 0.0 | 1 | p=1.00 |
| Base | Qwen-8B | Qwen-32B | 0.2 | 0.0 | -1.0 | inf | p=1.00 |

| | | | | | | | |
|---|---|---|---|---|---|---|---|
| Age shift | GPT-4.1 | GPT-4.1-nano | 0.0 | 1.8 | 1.0 | 0.0 | **p=0.004** |
| Age shift | GPT-4.1 | GPT-5 | 0.0 | 0.0 | 0.0 | 1 | p=1.00 |
| Age shift | GPT-4.1 | Llama-3-8B | 0.0 | 0.0 | 0.0 | 1 | p=1.00 |
| Age shift | GPT-4.1 | Qwen-8B | 0.0 | 0.2 | 1.0 | 0.0 | p=1.00 |
| Age shift | GPT-4.1 | Qwen-32B | 0.0 | 0.0 | 0.0 | 1 | p=1.00 |
| Age shift | GPT-4.1-nano | GPT-5 | 1.8 | 0.0 | -1.0 | inf | **p=0.004** |
| Age shift | GPT-4.1-nano | Llama-3-8B | 1.8 | 0.0 | -1.0 | inf | **p=0.004** |
| Age shift | GPT-4.1-nano | Qwen-8B | 1.8 | 0.2 | -0.889 | 9.147 | **p=0.021** |
| Age shift | GPT-4.1-nano | Qwen-32B | 1.8 | 0.0 | -1.0 | inf | **p=0.004** |
| Age shift | GPT-5 | Llama-3-8B | 0.0 | 0.0 | 0.0 | 1 | p=1.00 |
| Age shift | GPT-5 | Qwen-8B | 0.0 | 0.2 | 1.0 | 0.0 | p=1.00 |
| Age shift | GPT-5 | Qwen-32B | 0.0 | 0.0 | 0.0 | 1 | p=1.00 |
| Age shift | Llama-3-8B | Qwen-8B | 0.0 | 0.2 | 1.0 | 0.0 | p=1.00 |
| Age shift | Llama-3-8B | Qwen-32B | 0.0 | 0.0 | 0.0 | 1 | p=1.00 |
| Age shift | Qwen-8B | Qwen-32B | 0.2 | 0.0 | -1.0 | inf | p=1.00 |
| Header swap | GPT-4.1 | GPT-4.1-nano | 17.4 | 5.6 | -0.678 | 3.551 | **p<0.001** |
| Header swap | GPT-4.1 | GPT-5 | 17.4 | 0.0 | -1.0 | inf | **p<0.001** |
| Header swap | GPT-4.1 | Llama-3-8B | 17.4 | 0.0 | -1.0 | inf | **p<0.001** |

| | | | | | | | |
|---|---|---|---|---|---|---|---|
| Header swap | GPT-4.1 | Qwen-8B | 17.4 | 0.2 | -0.989 | 105.116 | **p<0.001** |
| Header swap | GPT-4.1 | Qwen-32B | 17.4 | 0.0 | -1.0 | inf | **p<0.001** |
| Header swap | GPT-4.1-nano | GPT-5 | 5.6 | 0.0 | -1.0 | inf | **p<0.001** |
| Header swap | GPT-4.1-nano | Llama-3-8B | 5.6 | 0.0 | -1.0 | inf | **p<0.001** |
| Header swap | GPT-4.1-nano | Qwen-8B | 5.6 | 0.2 | -0.964 | 29.602 | **p<0.001** |
| Header swap | GPT-4.1-nano | Qwen-32B | 5.6 | 0.0 | -1.0 | inf | **p<0.001** |
| Header swap | GPT-5 | Llama-3-8B | 0.0 | 0.0 | 0.0 | 1 | p=1.00 |
| Header swap | GPT-5 | Qwen-8B | 0.0 | 0.2 | 1.0 | 0.0 | p=1.00 |
| Header swap | GPT-5 | Qwen-32B | 0.0 | 0.0 | 0.0 | | p=1.00 |
| Header swap | Llama-3-8B | Qwen-8B | 0.0 | 0.2 | 1.0 | 0.0 | p=1.00 |
| Header swap | Llama-3-8B | Qwen-32B | 0.0 | 0.0 | 0.0 | | p=1.00 |
| Header swap | Qwen-8B | Qwen-32B | 0.2 | 0.0 | -1.0 | inf | p=1.00 |
| MRN digit change | GPT-4.1 | GPT-4.1-nano | 0.0 | 2.0 | 1.0 | 0.0 | p=0.002 |
| MRN digit change | GPT-4.1 | GPT-5 | 0.0 | 0.0 | 0.0 | | p=1.00 |
| MRN digit change | GPT-4.1 | Llama-3-8B | 0.0 | 0.0 | 0.0 | | p=1.00 |
| MRN digit change | GPT-4.1 | Qwen-8B | 0.0 | 0.2 | 1.0 | 0.0 | p=1.00 |
| MRN digit change | GPT-4.1 | Qwen-32B | 0.0 | 0.0 | 0.0 | | p=1.00 |
| MRN digit change | GPT-4.1-nano | GPT-5 | 2.0 | 0.0 | -1.0 | inf | p=0.002 |

| | | | | | | | |
|---|---|---|---|---|---|---|---|
| MRN digit change | GPT-4.1-nano | Llama-3-8B | 2.0 | 0.0 | -1.0 | inf | p=0.002 |
| MRN digit change | GPT-4.1-nano | Qwen-8B | 2.0 | 0.2 | -0.9 | 10.184 | p=0.011 |
| MRN digit change | GPT-4.1-nano | Qwen-32B | 2.0 | 0.0 | -1.0 | inf | p=0.002 |
| MRN digit change | GPT-5 | Llama-3-8B | 0.0 | 0.0 | 0.0 | | p=1.00 |
| MRN digit change | GPT-5 | Qwen-8B | 0.0 | 0.2 | 1.0 | 0.0 | p=1.00 |
| MRN digit change | GPT-5 | Qwen-32B | 0.0 | 0.0 | 0.0 | | p=1.00 |
| MRN digit change | Llama-3-8B | Qwen-8B | 0.0 | 0.2 | 1.0 | 0.0 | p=1.00 |
| MRN digit change | Llama-3-8B | Qwen-32B | 0.0 | 0.0 | 0.0 | | p=1.00 |
| MRN digit change | Qwen-8B | Qwen-32B | 0.2 | 0.0 | -1.0 | inf | p=1.00 |

**Supplementary Table 7** Token count per LLM model

| Model | Condition | Tokens Median | Total Tokens | Percents of the total |
|---|---|---|---|---|
| GPT-4.1 | Base (unmodified) | 271 | 135,500 | 6.4% |
| GPT-4.1 | Age shift | 271 | 135,500 | 6.4% |
| GPT-4.1 | Header swap | 271 | 135,500 | 6.4% |
| GPT-4.1 | MRN digit change | 271 | 135,500 | 6.4% |
| GPT-4.1-nano | Base (unmodified) | 288 | 144,000 | 6.8% |
| GPT-4.1-nano | Age shift | 286 | 143,000 | 6.8% |
| GPT-4.1-nano | Header swap | 286 | 143,000 | 6.8% |
| GPT-4.1-nano | MRN digit change | 307 | 153,500 | 7.3% |
| GPT-5 | Base (unmodified) | 296 | 148,000 | 7.0% |
| GPT-5 | Age shift | 296 | 148,000 | 7.0% |
| GPT-5 | Header swap | 296 | 148,000 | 7.0% |
| GPT-5 | MRN digit change | 296 | 148,000 | 7.0% |
| Llama-3-8B | Base (unmodified) | 82 | 41,000 | 1.9% |
| Llama-3-8B | Age shift | 82 | 41,000 | 1.9% |
| Llama-3-8B | Header swap | 82 | 41,000 | 1.9% |
| Llama-3-8B | MRN digit change | 82 | 41,000 | 1.9% |
| Qwen-8B | Base (unmodified) | 25 | 12,500 | 0.6% |
| Qwen-8B | Age shift | 25 | 12,500 | 0.6% |
| Qwen-8B | Header swap | 25 | 12,500 | 0.6% |
| Qwen-8B | MRN digit change | 25 | 12,500 | 0.6% |
| Qwen-32B | Base (unmodified) | 92 | 46,000 | 2.2% |
| Qwen-32B | Age shift | 92 | 46,000 | 2.2% |
| Qwen-32B | Header swap | 92 | 46,000 | 2.2% |
| Qwen-32B | MRN digit change | 92 | 46,000 | 2.2% |
| Commercial (ChatGPT) Subtotal | | — | 1,717,500 | 81.2% |
| Open-Source Subtotal | | — | 398,000 | 18.8% |
| GRAND TOTAL | | — | 2,115,500 | 100.0% |

**Supplementary Table 8** Token count pairwise comparison of different perturbations

| Model | Comparison | Base Tokens | Tamper Tokens | p-value |
|---|---|---|---|---|
| GPT-4.1 | Age shift vs Base | 271 | 271 | 1.0 |
| GPT-4.1 | Header swap vs Base | 271 | 271 | 1.0 |
| GPT-4.1 | MRN digit change vs Base | 271 | 271 | 1.0 |
| GPT-4.1-nano | Age shift vs Base | 288 | 286 | 0.62 |
| GPT-4.1-nano | Header swap vs Base | 288 | 286 | 0.62 |
| GPT-4.1-nano | MRN digit change vs Base | 288 | 307 | **<0.001** |
| GPT-5 | Age shift vs Base | 296 | 296 | 1.0 |
| GPT-5 | Header swap vs Base | 296 | 296 | 1.0 |
| GPT-5 | MRN digit change vs Base | 296 | 296 | 1.0 |
| Llama-3-8B | Age shift vs Base | 82 | 82 | 1.0 |
| Llama-3-8B | Header swap vs Base | 82 | 82 | 1.0 |
| Llama-3-8B | MRN digit change vs Base | 82 | 82 | 1.0 |
| Qwen-8B | Age shift vs Base | 25 | 25 | 1.0 |
| Qwen-8B | Header swap vs Base | 25 | 25 | 1.0 |
| Qwen-8B | MRN digit change vs Base | 25 | 25 | 1.0 |
| Qwen-32B | Age shift vs Base | 92 | 92 | 1.0 |
| Qwen-32B | Header swap vs Base | 92 | 92 | 1.0 |
| Qwen-32B | MRN digit change vs Base | 92 | 92 | 1.0 |

**Supplementary Table 9** Token count pairwise comparison across different LLMs

| Condition | Model 1 | Model 2 | Model 1 Tokens | Model_2_Tokens | Rank Biserial R | Rank Biserial Interpretation | p-value |
|---|---|---|---|---|---|---|---|
| Base | GPT-4.1 | GPT-4.1-nano | 271 | 288 | 0.059 | Negligible | **<0.01** |
| Base | GPT-4.1 | GPT-5 | 271 | 296 | 0.084 | Negligible | **<0.01** |
| Base | GPT-4.1 | Llama-3-8B | 271 | 82 | -0.697 | Large | **<0.01** |
| Base | GPT-4.1 | Qwen-8B | 271 | 25 | -0.908 | Large | **<0.01** |
| Base | GPT-4.1 | Qwen-32B | 271 | 92 | -0.661 | Large | **<0.01** |
| Base | GPT-4.1-nano | GPT-5 | 288 | 296 | 0.027 | Negligible | 0.054 |
| Base | GPT-4.1-nano | Llama-3-8B | 288 | 82 | -0.715 | Large | **<0.01** |
| Base | GPT-4.1-nano | Qwen-8B | 288 | 25 | -0.913 | Large | **<0.01** |
| Base | GPT-4.1-nano | Qwen-32B | 288 | 92 | -0.681 | Large | **<0.01** |
| Base | GPT-5 | Llama-3-8B | 296 | 82 | -0.723 | Large | **<0.01** |
| Base | GPT-5 | Qwen-8B | 296 | 25 | -0.916 | Large | **<0.01** |
| Base | GPT-5 | Qwen-32B | 296 | 92 | -0.689 | Large | **<0.01** |
| Base | Llama-3-8B | Qwen-8B | 82 | 25 | -0.695 | Large | **<0.01** |
| Base | Llama-3-8B | Qwen-32B | 82 | 92 | 0.109 | Small | **<0.01** |
| Base | Qwen-8B | Qwen-32B | 25 | 92 | 0.728 | Large | **<0.01** |
| Age shift | GPT-4.1 | GPT-4.1-nano | 271 | 286 | 0.052 | Negligible | **<0.01** |
| Age shift | GPT-4.1 | GPT-5 | 271 | 296 | 0.084 | Negligible | **<0.01** |
| Age shift | GPT-4.1 | Llama-3-8B | 271 | 82 | -0.697 | Large | **<0.01** |
| Age shift | GPT-4.1 | Qwen-8B | 271 | 25 | -0.908 | Large | **<0.01** |

| | | | | | | | |
|---|---|---|---|---|---|---|---|
| Age shift | GPT-4.1 | Qwen-32B | 271 | 92 | -0.661 | Large | **<0.01** |
| Age shift | GPT-4.1-nano | GPT-5 | 286 | 296 | 0.034 | Negligible | 0.015 |
| Age shift | GPT-4.1-nano | Llama-3-8B | 286 | 82 | -0.713 | Large | **<0.01** |
| Age shift | GPT-4.1-nano | Qwen-8B | 286 | 25 | -0.913 | Large | **<0.01** |
| Age shift | GPT-4.1-nano | Qwen-32B | 286 | 92 | -0.678 | Large | **<0.01** |
| Age shift | GPT-5 | Llama-3-8B | 296 | 82 | -0.723 | Large | **<0.01** |
| Age shift | GPT-5 | Qwen-8B | 296 | 25 | -0.916 | Large | **<0.01** |
| Age shift | GPT-5 | Qwen-32B | 296 | 92 | -0.689 | Large | **<0.01** |
| Age shift | Llama-3-8B | Qwen-8B | 82 | 25 | -0.695 | Large | **<0.01** |
| Age shift | Llama-3-8B | Qwen-32B | 82 | 92 | 0.109 | Small | **<0.01** |
| Age shift | Qwen-8B | Qwen-32B | 25 | 92 | 0.728 | Large | **<0.01** |
| Header swap | GPT-4.1 | GPT-4.1-nano | 271 | 286 | 0.052 | Negligible | **<0.01** |
| Header swap | GPT-4.1 | GPT-5 | 271 | 296 | 0.084 | Negligible | **<0.01** |
| Header swap | GPT-4.1 | Llama-3-8B | 271 | 82 | -0.697 | Large | **<0.01** |
| Header swap | GPT-4.1 | Qwen-8B | 271 | 25 | -0.908 | Large | **<0.01** |
| Header swap | GPT-4.1 | Qwen-32B | 271 | 92 | -0.661 | Large | **<0.01** |
| Header swap | GPT-4.1-nano | GPT-5 | 286 | 296 | 0.034 | Negligible | **<0.01** |
| Header swap | GPT-4.1-nano | Llama-3-8B | 286 | 82 | -0.713 | Large | **<0.01** |
| Header swap | GPT-4.1-nano | Qwen-8B | 286 | 25 | -0.913 | Large | **<0.01** |

| | | | | | | | |
|---|---|---|---|---|---|---|---|
| Header swap | GPT-4.1-nano | Qwen-32B | 286 | 92 | -0.678 | Large | **<0.01** |
| Header swap | GPT-5 | Llama-3-8B | 296 | 82 | -0.723 | Large | **<0.01** |
| Header swap | GPT-5 | Qwen-8B | 296 | 25 | -0.916 | Large | **<0.01** |
| Header swap | GPT-5 | Qwen-32B | 296 | 92 | -0.689 | Large | **<0.01** |
| Header swap | Llama-3-8B | Qwen-8B | 82 | 25 | -0.695 | Large | **<0.01** |
| Header swap | Llama-3-8B | Qwen-32B | 82 | 92 | 0.109 | Small | **<0.01** |
| Header swap | Qwen-8B | Qwen-32B | 25 | 92 | 0.728 | Large | **<0.01** |
| MRN digit change | GPT-4.1 | GPT-4.1-nano | 271 | 307 | 0.117 | Small | **<0.01** |
| MRN digit change | GPT-4.1 | GPT-5 | 271 | 296 | 0.084 | Negligible | **<0.01** |
| MRN digit change | GPT-4.1 | Llama-3-8B | 271 | 82 | -0.697 | Large | **<0.01** |
| MRN digit change | GPT-4.1 | Qwen-8B | 271 | 25 | -0.908 | Large | **<0.01** |
| MRN digit change | GPT-4.1 | Qwen-32B | 271 | 92 | -0.661 | Large | **<0.01** |
| MRN digit change | GPT-4.1-nano | GPT-5 | 307 | 296 | -0.036 | Negligible | **0.01** |
| MRN digit change | GPT-4.1-nano | Llama-3-8B | 307 | 82 | -0.733 | Large | **<0.01** |
| MRN digit change | GPT-4.1-nano | Qwen-8B | 307 | 25 | -0.919 | Large | **<0.01** |
| MRN digit change | GPT-4.1-nano | Qwen-32B | 307 | 92 | -0.7 | Large | **<0.01** |
| MRN digit change | GPT-5 | Llama-3-8B | 296 | 82 | -0.723 | Large | **<0.01** |

| | | | | | | | |
|---|---|---|---|---|---|---|---|
| MRN digit change | GPT-5 | Qwen-8B | 296 | 25 | -0.916 | Large | **<0.01** |
| MRN digit change | GPT-5 | Qwen-32B | 296 | 92 | -0.689 | Large | **<0.01** |
| MRN digit change | Llama-3-8B | Qwen-8B | 82 | 25 | -0.695 | Large | **<0.01** |
| MRN digit change | Llama-3-8B | Qwen-32B | 82 | 92 | 0.109 | Small | **<0.01** |
| MRN digit change | Qwen-8B | Qwen-32B | 25 | 92 | 0.728 | Large | **<0.01** |

**Supplementary Table 10** Latency count per LLMs

| Model | Condition | Observations (n) | Latency (ms), median [IQR] |
|---|---|---|---|
| **GPT-4.1** | Base (unmodified) | 500 | 13518 [12240–15836] |
| **GPT-4.1** | Age shift | 500 | 12964 [12031–15166] |
| **GPT-4.1** | Header swap | 500 | 13142 [12034–15451] |
| **GPT-4.1** | MRN digit change | 500 | 13076 [11966–15202] |
| **GPT-4.1-nano** | Base (unmodified) | 500 | 4870 [3403–7917] |
| **GPT-4.1-nano** | Age shift | 500 | 6116 [3388–8100] |
| **GPT-4.1-nano** | Header swap | 500 | 5893 [3365–8424] |
| **GPT-4.1-nano** | MRN digit change | 500 | 4726 [3410–7948] |
| **GPT-5** | Base (unmodified) | 500 | 16332 [15350–17322] |
| **GPT-5** | Age shift | 500 | 16136 [15086–17168] |
| **GPT-5** | Header swap | 500 | 16038 [15013–17238] |
| **GPT-5** | MRN digit change | 500 | 16035 [15028–17196] |
| **Llama-3-8B** | Base (unmodified) | 500 | 31807 [31242–32562] |
| **Llama-3-8B** | Age shift | 500 | 32026 [31284–32556] |
| **Llama-3-8B** | Header swap | 500 | 32000 [31270–32510] |
| **Llama-3-8B** | MRN digit change | 500 | 31948 [31283–32533] |
| **Qwen-8B** | Base (unmodified) | 500 | 54004 [53449–54402] |
| **Qwen-8B** | Age shift | 500 | 54004 [53474–54392] |
| **Qwen-8B** | Header swap | 500 | 53996 [53444–54344] |
| **Qwen-8B** | MRN digit change | 500 | 53980 [53463–54368] |
| **Qwen-32B** | Base (unmodified) | 500 | 121813 [119472–122934] |
| **Qwen-32B** | Age shift | 500 | 122162 [119566–122909] |
| **Qwen-32B** | Header swap | 500 | 122044 [119579–122899] |
| **Qwen-32B** | MRN digit change | 500 | 122014 [119464–122983] |

**Supplementary Table 11** Latency count pairwise comparison across different LLMs

| Condition | Comparison | Model 1 Median (ms) | Model 2 Median (ms) | p-value | Rank-biserial r |
|---|---|---|---|---|---|
| **Base (unmodified)** | GPT-4.1 vs GPT-5 | 13518 | 16332 | **<0.001** | 0.413 |
| **Base (unmodified)** | GPT-4.1 vs GPT-4.1-nano | 13518 | 4870 | **<0.001** | -0.959 |
| **Base (unmodified)** | GPT-4.1 vs Llama-3-8B | 13518 | 31807 | **<0.001** | 0.962 |
| **Base (unmodified)** | GPT-4.1 vs Qwen-32B | 13518 | 121813 | **<0.001** | 1 |
| **Base (unmodified)** | GPT-4.1 vs Qwen-8B | 13518 | 54004 | **<0.001** | 0.988 |
| **Base (unmodified)** | GPT-5 vs GPT-4.1-nano | 16332 | 4870 | **<0.001** | -0.785 |
| **Base (unmodified)** | GPT-5 vs Llama-3-8B | 16332 | 31807 | **<0.001** | 1 |
| **Base (unmodified)** | GPT-5 vs Qwen-32B | 16332 | 121813 | **<0.001** | 1 |
| **Base (unmodified)** | GPT-5 vs Qwen-8B | 16332 | 54004 | **<0.001** | 1 |
| **Base (unmodified)** | GPT-4.1-nano vs Llama-3-8B | 4870 | 31807 | **<0.001** | 1 |
| **Base (unmodified)** | GPT-4.1-nano vs Qwen-32B | 4870 | 121813 | **<0.001** | 1 |
| **Base (unmodified)** | GPT-4.1-nano vs Qwen-8B | 4870 | 54004 | **<0.001** | 1 |
| **Base (unmodified)** | Llama-3-8B vs Qwen-32B | 31807 | 121813 | **<0.001** | 0.964 |
| **Base (unmodified)** | Llama-3-8B vs Qwen-8B | 31807 | 54004 | **<0.001** | 0.964 |
| **Base (unmodified)** | Qwen-32B vs Qwen-8B | 121813 | 54004 | **<0.001** | -1 |
| **Age shift** | GPT-4.1 vs GPT-5 | 12964 | 16136 | **<0.001** | 0.436 |
| **Age shift** | GPT-4.1 vs GPT-4.1-nano | 12964 | 6116 | **<0.001** | -0.926 |
| **Age shift** | GPT-4.1 vs Llama-3-8B | 12964 | 32026 | **<0.001** | 0.982 |
| **Age shift** | GPT-4.1 vs Qwen-32B | 12964 | 122162 | **<0.001** | 1 |
| **Age shift** | GPT-4.1 vs Qwen-8B | 12964 | 54004 | **<0.001** | 0.996 |
| **Age shift** | GPT-5 vs GPT-4.1-nano | 16136 | 6116 | **<0.001** | -0.697 |
| **Age shift** | GPT-5 vs Llama-3-8B | 16136 | 32026 | **<0.001** | 1 |
| **Age shift** | GPT-5 vs Qwen-32B | 16136 | 122162 | **<0.001** | 1 |
| **Age shift** | GPT-5 vs Qwen-8B | 16136 | 54004 | **<0.001** | 1 |
| **Age shift** | GPT-4.1-nano vs Llama-3-8B | 6116 | 32026 | **<0.001** | 0.996 |
| **Age shift** | GPT-4.1-nano vs Qwen-32B | 6116 | 122162 | **<0.001** | 1 |
| **Age shift** | GPT-4.1-nano vs Qwen-8B | 6116 | 54004 | **<0.001** | 0.996 |
| **Age shift** | Llama-3-8B vs Qwen-32B | 32026 | 122162 | **<0.001** | 0.964 |
| **Age shift** | Llama-3-8B vs Qwen-8B | 32026 | 54004 | **<0.001** | 0.964 |
| **Age shift** | Qwen-32B vs Qwen-8B | 122162 | 54004 | **<0.001** | -1 |
| **Header swap** | GPT-4.1 vs GPT-5 | 13142 | 16038 | **<0.001** | 0.406 |

| | | | | | |
|---|---|---|---|---|---|
| **Header swap** | GPT-4.1 vs GPT-4.1-nano | 13142 | 5893 | **<0.001** | -0.947 |
| **Header swap** | GPT-4.1 vs Llama-3-8B | 13142 | 32000 | **<0.001** | 0.982 |
| **Header swap** | GPT-4.1 vs Qwen-32B | 13142 | 122044 | **<0.001** | 1 |
| **Header swap** | GPT-4.1 vs Qwen-8B | 13142 | 53996 | **<0.001** | 1 |
| **Header swap** | GPT-5 vs GPT-4.1-nano | 16038 | 5893 | **<0.001** | -0.721 |
| **Header swap** | GPT-5 vs Llama-3-8B | 16038 | 32000 | **<0.001** | 0.996 |
| **Header swap** | GPT-5 vs Qwen-32B | 16038 | 122044 | **<0.001** | 1 |
| **Header swap** | GPT-5 vs Qwen-8B | 16038 | 53996 | **<0.001** | 1 |
| **Header swap** | GPT-4.1-nano vs Llama-3-8B | 5893 | 32000 | **<0.001** | 1 |
| **Header swap** | GPT-4.1-nano vs Qwen-32B | 5893 | 122044 | **<0.001** | 1 |
| **Header swap** | GPT-4.1-nano vs Qwen-8B | 5893 | 53996 | **<0.001** | 1 |
| **Header swap** | Llama-3-8B vs Qwen-32B | 32000 | 122044 | **<0.001** | 0.964 |
| **Header swap** | Llama-3-8B vs Qwen-8B | 32000 | 53996 | **<0.001** | 0.964 |
| **Header swap** | Qwen-32B vs Qwen-8B | 122044 | 53996 | **<0.001** | -1 |
| **MRN digit change** | GPT-4.1 vs GPT-5 | 13076 | 16035 | **<0.001** | 0.377 |
| **MRN digit change** | GPT-4.1 vs GPT-4.1-nano | 13076 | 4726 | **<0.001** | -0.917 |
| **MRN digit change** | GPT-4.1 vs Llama-3-8B | 13076 | 31948 | **<0.001** | 0.976 |
| **MRN digit change** | GPT-4.1 vs Qwen-32B | 13076 | 122014 | **<0.001** | 0.992 |
| **MRN digit change** | GPT-4.1 vs Qwen-8B | 13076 | 53980 | **<0.001** | 0.992 |
| **MRN digit change** | GPT-5 vs GPT-4.1-nano | 16035 | 4726 | **<0.001** | -0.639 |
| **MRN digit change** | GPT-5 vs Llama-3-8B | 16035 | 31948 | **<0.001** | 1 |
| **MRN digit change** | GPT-5 vs Qwen-32B | 16035 | 122014 | **<0.001** | 1 |
| **MRN digit change** | GPT-5 vs Qwen-8B | 16035 | 53980 | **<0.001** | 1 |
| **MRN digit change** | GPT-4.1-nano vs Llama-3-8B | 4726 | 31948 | **<0.001** | 0.996 |
| **MRN digit change** | GPT-4.1-nano vs Qwen-32B | 4726 | 122014 | **<0.001** | 0.996 |
| **MRN digit change** | GPT-4.1-nano vs Qwen-8B | 4726 | 53980 | **<0.001** | 0.996 |
| **MRN digit change** | Llama-3-8B vs Qwen-32B | 31948 | 122014 | **<0.001** | 0.964 |
| **MRN digit change** | Llama-3-8B vs Qwen-8B | 31948 | 53980 | **<0.001** | 0.964 |
| **MRN digit change** | Qwen-32B vs Qwen-8B | 122014 | 53980 | **<0.001** | -1 |

Division of Data-Driven and Digital Medicine (D3M), Icahn School of Medicine at Mount Sinai, New York, USA
